# Investigating the Experiences of Frontline Health Workers in the Delivery of Healthcare during the COVID-19 Pandemic in Ghana

**DOI:** 10.1101/2025.11.22.25338669

**Authors:** Faustina Hayford Blankson, Nathaniel Larbi Andah, Hannah Brifo

**Author notes:** Corresponding author: (FHB). These authors contributed equally to this work.

## Abstract

The COVID-19 pandemic has had a profound impact on the lives of many in Ghana and globally, including the frontline health workers (FHWs). FHWs provide diverse services, including treatment, care, contact tracing, and public education and awareness during the pandemic. Most of these workers had direct daily contact and interactions with people infected with the virus. However, their experiences, challenges, strategies and motivations as they performed their duties have not been thoroughly examined, particularly in Ghana.

**Objective:** This study examined the experiences of FHWs of COVID-19 in Ghana in order to understand their challenges, strategies and motivations for their work. The results of this study can be used to plan and implement appropriate policies, strategies and interventions in future health emergencies.

**Method:** The study employed primarily a qualitative design with an interpretive approach even though in some cases frequencies and percentages were reported to compliment the narrative responses. A purposive sampling procedure was used. Delphi technique served as the primary data collection technique. Participants for this study were thirty-two (32) FHWs drawn from three regions of Ghana; Greater Accra, Ashanti, and Western regions, from 20^th^ August 2022 to 22^nd^ February 2023.

**Results:** The results of the study revealed that FHWs faced several challenges during the COVID-19 pandemic. These include high-stress levels, inadequate resources, patients’ spiritual beliefs about the disease, burnout due to work overload, inadequate support system, bribery by people who test positive, and difficulty in balancing family life and work. They employed different strategies to mitigate these challenges, including taking adequate rest, self-motivation, and satisfaction from provision of accurate information to patients, maintaining calm, self-control and assuring patients. Finally, they were motivated to perform their daily tasks by the incentive and reward packages instituted by the government as well as satisfaction they got from providing high-quality care and treatment to patients.

**Conclusion:** The study concludes that because FHWs play a pivotal role in pandemic emergencies, their working conditions, health and wellbeing must be taken seriously to enable them provide the needed services for patients. This study adds to the body of knowledge on the subject matter, provides insight to ease the burden of FHWs during emergencies, provides information to guide policy and decision making, and creates a basis for future research.

## Introduction

Frontline healthcare workers (FHWs) faced extraordinary challenges in the wake of the COVID-19 pandemic (Li & Luo, 2020). COVID-19, a communicable disease caused by a new coronavirus, which rapidly spread across the globe (World Health Organization, 2020). These healthcare professionals faced both physical risks and psychological stress as they bear the brunt of the outbreak (Xiang et al., 2020; Lance et al., 2020). They also grappled with an overwhelming workload and a shortage of essential personal protective equipment (PPE) (Li & Luo, 2020; Cherish, Gray, Fairlie et al., 2020). Additionally, the pandemic generated widespread fear and anxiety in the general population, further straining healthcare workers who were at the forefront of providing care (Duncan, Schaller & Park, 2015; Pappas et al., 2016). The COVID-19 pandemic revealed critical vulnerabilities within the healthcare system, necessitating urgent support and resources to safeguard the well-being of these dedicated professionals (Stuijfzand et al., 2020). While several studies have explored the experiences of FHWs during the COVID-19 pandemic worldwide, limited attention has been given to the situation in developing societies like Ghana, with even fewer studies taking a qualitative approach and considering the narrative experiences of the health workers. It is crucial to gain insight into the daily experiences, challenges, coping strategies and motivations of the FHWs, as their well-being is closely tied to their cognitive functioning and job performance (Husain et al., 2020; Lee et al., 2018). This study was, therefore, designed to examine the holistic experiences of the frontline health workers in Ghana so that appropriate policies and interventions will be developed to address the gaps observed in preparation for future health emergencies.

### Theoretical Framework: The Social Ecological Model

The study was guided by the Social Ecological Model. This model and its multi-faceted approach to influences provide a framework for gaining a deeper insight into the frontline health workers experiences and challenges in providing effective and efficient services, as well as understanding their coping mechanisms utilized during those challenging times of COVID-19 pandemic. *Below is an illustration of the Social Ecological Model*

**Fig 1:**
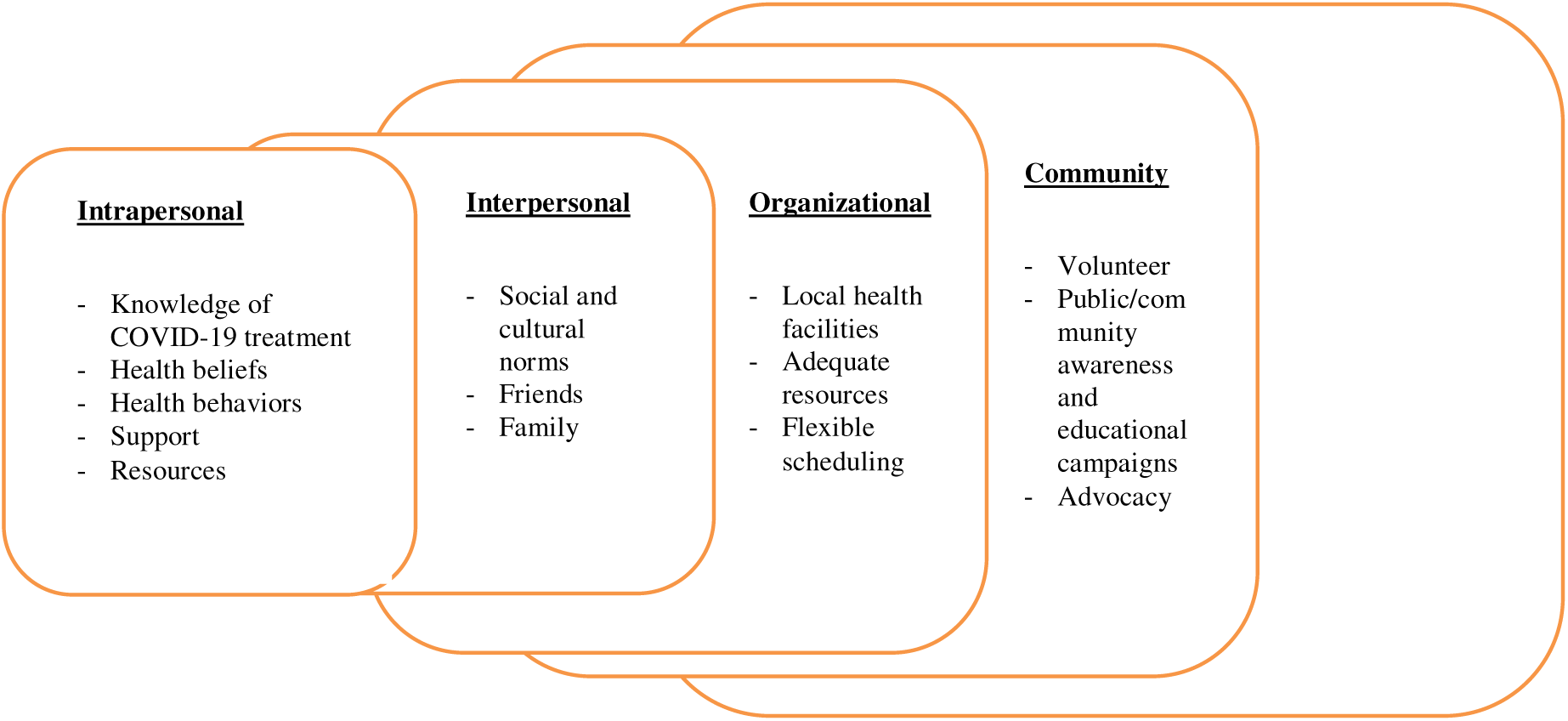
Illustration of the Social Ecological Model (SEM) Adapted from Blankson, 2017

The following explains how the researchers applied the model to the study.

#### Intrapersonal and Interpersonal

The first two levels of the model enabled the researchers to obtain information about participants’ beliefs, knowledge, resources, support, and attitudes regarding health care services provided to COVID-19 patients. Attention was also paid to the FHWs’ social and cultural norms; family belief systems and their family communication processes in relation to their contributions to the fight of COVID-19.

#### Institutional and Community

At the institutional and community levels, the participants were asked to identify the institutional challenges they faced when treating COVID-19 patients.

#### Policy

Health policy level information was not solicited because it was not within the scope of this study. Although this study did not focus on policy, participants’ responses indicated an important policy which is an insurance package with an assured sum of GHc350, 000 (approx. US$29,411) created by policy makers during the pandemic but did not materialize.

## Method

### Study design

The study was primarily qualitative in design with a combination of social constructivist and interpretivist paradigms. In certain instances, numerical data in the form of simple frequencies and percentages were obtained to support or corroborate the narrative responses of participants (Creswell, 2018). The qualitative research design was appropriate because this research primarily aimed at exploring and interpreting the daily experiences of FHWs during the COVID-19 pandemic in Ghana.

### Inclusion and exclusion criteria

Frontline health workers with responsibility for any corona virus-related work were eligible to participate in the study. Health workers not assigned to perform a corona virus related work were excluded from the study. Other health workers who were not in a position to adequately answer the questions were also excluded.

### Sampling method

The purposive sampling technique was used to select 32 participants for the study. As a nonprobability sampling method, the purposive technique was used to recruit participants because of the uniqueness of the perspectives and research questions being investigated in the study.

### Data collection

Data were collected from 20^th^ August 2022 to 22^nd^ February 2023 in three regions of Ghana (Western, Ashanti, and Greater Accra). These regions were selected because they had the highest case count of the Coronavirus (COVID-19) in the nation (Ghana Health Service, 2021). Data were collected from thirty two (32) FHWs with 10 each from Western and Greater Accra regions and 12 from the Ashanti region. An interview guide was used to collect data on the experiences, challenges, strategies and motivations of the FHWs during the COVID-19 pandemic. Participants’ socio-demographic information was collected to provide contextual and background information. The guide was created using Google Forms and consisted of both closed- and open-ended questions aimed at soliciting in-depth information from participants. The Delphi technique was used as the main data collection technique. This allowed the researchers to collect in-depth interview data via Google Forms online tool from participants who did not have time for a face-to-face or focus group interview (Fertman & Allensworth, 2017).

### Data processing and analysis

The qualitative data collected were meticulously reviewed and transcribed after each day’s interviews. The principal investigator transcribed and analysed the interview responses, utilizing both inductive and deductive approaches of analysis. The deductive approach was utilized to group the data and examine relationships, while the inductive approach was done using the research questions. The data were analysed using Microsoft Excel. The narrative statements are presented as quotations while the demographic and numerical results are presented as simple frequencies and percentages using tables where applicable. The data were categorized by region, color coded, and analyzed. Themes were developed to ensure a commonality of groupings and to enhance understanding of the experiences of the FHWs. Questions that were incorrectly answered were returned for clarity.

The following research questions guided the study:

1. What were the experiences of FHWs as they provide care for COVID-19 patients in Ghana?
2. What challenges did the FHWs face as they perform their duties?
3. What strategies did the FHWs employ to mitigate the challenges they face at work?
4. What motivated FHWs to perform their daily tasks during the pandemic?

### Confidential and Ethical Considerations

Approval for this study was obtained from the Ethics Review Committee of the Ghana Health Service (GHS) with approval number **GHS-ERC:013/04/22**. All participants were informed about the study’s purpose, procedures, potential risks, and benefits before data collection. Participation was voluntary, and participants were free to withdraw at any stage without any penalty.

Informed written consent was obtained from all participants before the interview questions were shared. The consent form was sent to each participant via email and electronically signed and returned to the research team before participation.

Confidentiality was maintained by using coded identification numbers instead of names. Data were stored on password-protected devices accessible only to the research team. All identifying information was removed during transcription and analysis. The data was securely stored for three years and then deleted. This study posed no physical or psychological risk to participants.

## Results

### Characteristics of Participants

The characteristics of the 32 FHWs interviewed for the study is presented in Table 1 below. The participants range from ages 25 to 60 with the majority (56%) between ages 30 and 49. Those in their 20s constituted 28% and those 50 years and above formed 16%. There was a balance in gender representation, with 47% females and 53% males. Fifty-nine percent (59%) were married compared to 34% single and 2% divorced or widowed. Most of the participants were physicians (31%), followed by nurses and lab assistants (28% each), and auxiliary staff (13%). Participants were from government hospitals (37.5%), government clinics (21.9%), private hospitals (25%), and private clinics (15.6%). With regard to participants’ duties as frontline health workers, 56% provided services to patients at the hospitals, clinics and community centres, 28% work at the quarantine or isolation centres, and 13% engaged in testing for the virus. The participants were evenly selected from the three regions, 10 each from Western and Greater Accra and 12 from Ashanti region. In terms of gender distribution, there were no significant differences for all the participants’ demographic characteristics.

**Table 1:** Characteristics of study participants (n=32)

| Characteristics of Participants | Total<br>(Percentage) | By Gender |  |
| --- | --- | --- | --- |
|  |  | Male | Female |
| **Age** |  |  |  |
| 20s | 9 (28%) | 5 | 4 |
| 30s | 9 (28%) | 4 | 5 |
| 40s | 9 (28%) | 4 | 5 |
| 50s | 5 (16%) | 4 | 1 |
| 60s | 0 | 0 | 0 |

| Gender |  |
| --- | --- |
| Female | 15 (47%) |
| Male | 17 (53%) |

### Thematic Analysis of Participants’ Responses

The responses from the 32 FHWs interviewed were analysed in line with the study’s research questions. The themes that emerged are presented as follows.

### RQ1: Experiences of FHWs in the performance of their duties

Research question one solicited the experiences of the FHWs as they performed their tasks during the COVID-19 pandemic in Ghana. Dominant themes that emerged under this included feeling of anxiety and fear while caring for patients, treatment of first COVID-19 case, training of health professionals, and adherence to strict treatment protocols and decisions.

#### Anxiety and fear when caring for patients

Participants were first asked to indicate what their reaction was when they first learned that they will be caring for COVID-19 patients. Their responses varied from fear and anxiety to excitement and normal feeling. The majority of the FHWs interviewed (63%) stated that they were afraid, scared and anxious when told to care for or treat COVID-19 patients, despite having a good knowledge about the virus, its causes, prevalence, and other behavioral characteristics. They attributed their fear and anxiety to the high incidence of death among the FHWs. Despite acknowledging their anxiety, they believed and assured themselves that they could only be infected if they ignored to follow safety protocols, as prescribed by the World Health Organization. Some of the FHWs indicated they became frightened after watching circulating videos of COVID-19 patients on social media. A few of them related their fear and anxiety to losing close relatives and friends. The excerpts below reflect some of the participants’ feelings.

> *I got very scared initially but as a Christian, I prayed about it and I discussed the news of going to the COVID Center with my wife. We prayed together as a couple and I got encouraged to go and save lives (Male, 30’s).*
>
> *I was scared because of how I lost a friend to this COVID-19 (Female, 40’s,).*
>
> *I was anxious and scared at the same time but one thing that kept me going was the fact that if I follow the safety protocols laid down by WHO, I won’t be infected (Female, 30’s).*

On the contrary, 6 participants (19%) indicated they were not worried or afraid but rather curious and had to show a brave face to offer care or treatment to the patients. Asked how they felt, they stated, *Excited (Male, 50’s)*.

> *I was prepared to attend to my patients. I was not afraid (Female, 30’s) Curious. But I showed a brave face. I was so much careful (Male, 30’s).*
>
> *My reaction was normal. Since I had passion in my field of work, I had to work to save a life (Male, 50’s).*

### Treatment of first COVID-19 cases

The participants were also asked to share their experience when they received and treated their first COVID-19 patient. The study result showed that that majority of the FHWs experienced mixed emotions the first time they treated patients with COVID-19 symptoms. While many indicated that they were scared or terrified (14 participants; 44%), very few (4 participants; 12%) responded they were calm or felt normal. Others were apprehensive, felt bad, sad, worried or pity for the patients. The sentiment expressed by a female captured this mixed emotion.

> *When I saw the patient, I felt sad and pity for her. I was eager to start treatment (Female, 20’s).*

Many of the FHWs indicated they sought guidance from their supervisors when they had their first COVID-19 cases. Only a handful admitted to not seeking guidance but rather adopted a cautious approach to treating their patients. Some of the FHWs acknowledged they had difficulties wearing the personal protective equipment (PPE) for the first time. The statements below capture some of their encounters with their first COVID-19 cases.

> *I was surprised that truly people have indeed been infected with the virus (Female, 40’s).*
>
> *I was scared that I might not be cautious enough and I’ll get infected in their care (Male, 30’s).*
>
> *My team leaders were present at the facility before the first batch of cases were brought into the facility so they* assisted in the process of receiving the patient (Male, 50’s).

#### Preparation and readiness

The participants were asked about how prepared they were to treat or care for COVID-19 patients. Their responses indicate a high level of preparation and readiness, with only 5 participants (16%) stating that they were not prepared to handle the cases or provide care to patients. The majority (74%) stated that they were adequately, well prepared or very prepared and ready to treat or care for their patients. They cited their training for emergency cases in infection prevention and control and following safety protocols as key factors. In addition, they identified the initial availability of resources such as PPEs, N95 masks, gloves, and sanitizers to aid in patient treatment. Similarly, the participants believed that emphasis given to public education by health institutions and the government on safety guidelines and protocols during the pandemic eased their anxiety. Below are some of the sentiments expressed by three interviewees.

> *The hospital organized refresher courses for staff so there was enough knowledge on how to care for COVID-19 patients (Female, 20’s).*
>
> *Provision of PPEs were given and also enough education on the condition has been provided already. So, I had the skills on what to wear, what not to do and what to do (Female, 30’s).*
>
> *Knowing how dangerous and deadly covid-19 is, I personally psyched myself to defy all odds to save lives. More importantly, we were trained as a team in infection prevention and control (IPC) in addition to all skills relevant in managing COVID cases (Female, 30’s).*

#### Treatment decisions

The participants were asked to share how they made decisions on caring for or treatment of patients with COVID-19. A number of factors were mentioned, including the severity of the patient’s condition, adherence to case management protocols, reliance on assessment reports, laboratory results, and guidelines developed by the National Case Management Team and World Health Organization (WHO). The majority of the participants indicated their decisions were guided by the severity of the case (64%) and strict case management protocols (60%). Only a few sought advice from colleagues handling COVID-19 cases. The statements below explain how treatment decisions were made by some of the health workers.

> *Treatment decision is made based on the severity and the case management protocols. As a member of the National Case Management team, we had developed protocols for case management (Male, 40’s).*
>
> *I assess all cases coming in and depending on the severity of some the needed treatment is given accordingly (Female, 30’s).*
>
> *Treatment is mainly based on assessment, history taking, and laboratory test to provide adequate and exact holistic care for patients with COVID-19 (Female, 40’s).*

### RQ2. Pressures and Challenges faced when treating and caring for COVID-19 patients

Research question two sought to understand the pressures and challenges the FHWs faced in performing their duties. Two questions guided this section of the interview. First, the participants were asked to share the pressures they encountered in their work. They were then asked to indicate the challenges that hindered their ability to treat or care for their COVID-19 patients. Their responses are shared below.

#### Pressures faced at work

The participants indicated facing a number of pressures as FHWs. Eleven participants (34%) identified lack of cooperation or non-compliance by patients as the major pressure they faced. This was followed by pressure from not having adequate resources, equipment or logistics to work with (23%), pressure and demands from patients, families and friends of the patients (16%), delays in receiving lab results (9%), and dealing with large number of patients (9%). Participants who identified pressure emanating from lack of cooperation or non-compliance by patients supported their claims with the following statements.

> *Some patients were resisting confinement and we had to use any ways possible to let them understand that they have to be isolated to stop the spread of the virus (Female, 30’s).*
>
> *Patients not properly isolating brought a lot of pressure on us because you have to be checking and monitoring them in addition to the work you are doing. They didn’t believe they had the condition and so were not cooperating at all (Male, 40’s).*
>
> *Strong denials by patients that they do not have COVID. Checking vital signs for sometimes over 100 patients and preparing them for doctors’ review (Female, 30’s).*

The statement below by a male physician represents sentiments expressed by the FHWs who identified pressure from not having adequate resources, equipment or logistics to work with. He stated,

> *Limited resources brought pressure on us, for instance when we had only one flow meter for oxygenation which was being used, yet we had a COVID case that equally needs oxygenation and now you have to prioritize who needs it more or better still move to other departments for a flow meter Secondly the referral points almost always alleged they had no beds so the polyclinic had to keep patients and manage for days or even discharge (Male, 30’s).*

Participants who experienced pressure and demands from patients, families and friends of their patients supported their claims with the following statements.

> *The demand by patients and their relatives for home treatment was too much. Especially patient’s relatives wanting to enter and be in the isolation room with their loved ones was also challenging to us and it compounded to the pressure we are facing (Male, 30’s).*
>
> *Patients and their families always demanding that we speak to the government to pay compensations to them as they claim we the staff are only helping the government to make money by keeping them in isolation (Female, 50’s).*

#### Challenges faced by frontline health workers

The study participants were also asked to identify the challenges they faced as FHWs caring for and treating COVID-19 patients. Their responses are presented.

##### Lack of resources

Nearly all the FHWs interviewed (87%) identified lack of resources, equipment or logistics as their major challenge in providing care to patients with COVID-19. They complained of shortage of personal protective equipment, ventilators, flow meters, and vaccines in their health facilities. Three of the participants stated,

> *The lack of PPEs brought about the challenges I faced (Male, 40’s).*
>
> *Ventilators and flow meters were what we lacked in the hospital. It posed a lot of challenges to me and my team (Male, 40’s).*
>
> *Vaccines were also in shortage and there was an increase in hesitancy (Female, 30’s).*

Some participants went further to ascribe the inadequate resources to their feeling of unhappiness about their inability to provide the desired support for their COVID-19 patients. They expressed disappointment in the health facilities’ inability to provide them with the necessary resources to treat patients. The statements below represent sentiments expressed by these participants.

> *I get disappointed especially when I see them in crises and can’t do anything about it due to lack of resources (Female, 30’s).*
>
> *I mostly don’t feel happy if I am not able to help a patient. Most especially if the patient’s requests are beyond me as a nurse (Female, 40’s).*
>
> *Although we lacked a lot of things needed to provide care, my team and I improvised when necessary to still give standardized care to patients but it was still not enough and that makes me sad (Female, 40’s).*

##### Stress and burnout

Stress was identified by 18 FHWs (56%) as a major challenge. These participants attributed their stress to a number of factors including patient con-compliance to safety protocols and medications, inadequate resources to treat patients, high patient-to-physician ratio, inability to take necessary breaks to rest, and difficulty in managing their overwhelming daily workloads. Almost all of them complained of burnout. These statements were made to support the claims.

> *I have sleepless nights since most of the cases are brought in the night (Female, 40’s).*
>
> *I get so tired during the day because I do a lot of work of two people at the same time, this makes me burned (Female, 30’s).*
>
> *There is no time to rest because you are needed every day due to the high recorded cases in the hospital (Male, 40’s).*
>
> *I am responsible for caring for multiple critically ill patients, constantly adjusting ventilator settings, administering medications, and providing emotional support to families of patients. I was so stressed to the point that I struggled to maintain a sense of composure while witnessing the suffering and high mortality rates among my patients (Female, 30’s).*
>
> *As a physician, I worked at the emergency department during the COVID-19 surge. I was faced with limited ventilators and ICU beds forcing me to make heart-breaking decisions about which patients receive critical care. This was so stressful to me (Female, 40’s).*

#### Patient spiritual beliefs

Some participants identified patients’ spiritual beliefs about COVID-19 as a challenge. They complained that some patients refused to accept their positive test results and treatment because of spiritual reasons. These patients believe the COVID-19 causes to be spiritual, and so treatment should also be spiritual. Two of the FHWs interviewed captured their frustrations in these statements,

> *Most of the patients with spiritual beliefs are not cooperative at all. They turn to take anointing oil instead of medicine with the hope that God will protect them from the virus (Male, 30’s).*
>
> *Some patients believe that they are immune to the virus even when their lab results are positive. They refused to take treatment because of their spiritual beliefs (Female, 30’s).*

According to the FHWs interviewed, patients’ spiritual beliefs influenced their decisions to comply with treatment and follow safety protocols. Some patients had scepticism towards test results, leading to non-compliance with prescribed medications. Additionally, a significant proportion of COVID-19 patients in isolation centers prioritized spirituality over scientific causes of the disease, with prayer often being prioritized over adherence to treatment regimens. Three of the interviewees summarized the frustrations of FHWs in these statements,

> *Some of the patients are unwilling to cooperate with staff because they don’t believe the test is accurate to declare them positive for COVID-19 and hence the need to stay in isolation (Male, 40’s).*
>
> *The patients I came in contact with are all spiritual and nothing you say about the treatment regime makes sense to them (Female, 30’s).*
>
> *I realised that even though my patients were able to adhere to the safety protocols at the center, they placed God first in all things they did at the center (Male, 50’s).*

#### Inadequate staff

In addition to other challenges, 12 of the participants (37%) identified inadequate staffing as hindering their ability to offer effective care to patients. They complained that staff were required to perform double duties or shifts to compensate for the limited personnel. This, according to them, significantly impacted the quality of service they delivered to their patients. Some of the participants complained,

> *We lack staff at my place of work, so I have to be running a twelve-hour shift (Male, 30’s).*
>
> *Just imagine how I have to be going to different wards carrying out a treatment plan for patients and at the same time taking patient’s case history at the front desk (Female, 30’s).*
>
> *We have only a few staff in the hospital and the number of COVID-19 cases is more than the staff so it puts a lot of pressure on us making us burnout so easily (Male, 40’s).*
>
> *When the COVID-19 was high, most of my colleagues were sick as a result of stress and that made it hard for the rest of us to handle patients (Male, 50’s).*

#### Incentives to alter test results or avoid isolation

About 25% of the participants complained of attempts by patients, family members or friends to influence their COVID-19 test results or decision to quarantine or isolate them. According to these FHWs, the patients and their families or friends tried to offer monetary inducements to supervisors of the isolation centers to write them an exemption or clearance report. Others attempted to bribe physicians or lab technicians in exchange for positive test results. All the FHWs interviewed expressed their disgust over this aberrant behaviour. They offered the following statements to substantiate their claims.

> *Some patients want to pay money so I could discharge them from the isolation center (Male, 40’s).*
>
> *As a lab technician, I do get a lot of offers coming from patients who have tested positive for COVID-19. They offer to pay huge money for the lab results to be altered (Male, 30’s).*
>
> *Relatives of some of the COVID-19 patients will intentionally put money in an envelope when they come visiting. That money is there for me to do something, it is not free (Male, 30’s).*

#### Delayed lab results

A few participants (19%) complained of long delays in receiving lab results for the COVID-19 patients. According to them, this caused significant setbacks for both patients and physicians. Some of the patients in isolation centers were frustrated and anxious while waiting for confirmation of their results. Three of the participants had these to say,

> *The lab tests can take like forever for the test results to be out, this increases our stress level by way of compiling our workload (Male, 50’s).*
>
> *As a lab technician, it takes some few hours to get the lab results ready but due to the high number of blood samples the test results prolong (Male, 30’s).*
>
> *The truth is that the results of patients take longer to be ready not because there are no technicians but the equipment to carry it out is limited and there are a lot of them (Male, 30’s).*

#### Balancing family life and work

Another challenge faced by the FHWs interviewed was difficulty in balancing family life and work. At least 20% of the participants identified this as a major challenge. According to them, this was mainly because of complaints from their families, particularly because of the isolation and long periods they were away from their families. They resorted to the use of cell phones, social media, video calls and ZOOM to connect and communicate with their families. Three of the participants explained,

> *Leaving my children to visit my patients was so hard (Female, 30’s).*
>
> *I normally connect with my families through Zoom and sometimes video calls even though I am not able to go home frequently (Male, 50’s).*
>
> *I can go home at least twice a week to see my family, so balancing work and family life is a bit easier for me because of the support I get from my family (Female, 30’s).*

### RQ3: Strategies to Mitigate the Challenges FHWs Faced

The third research question asked the FHWs to share the strategies used to mitigate or minimize the challenges they faced. The themes that emerged from their responses are presented.

#### Adequate rest

Nearly all the FHWs interviewed identified resting after work as critical in their line of work. According to them, they often took long hours of rest when off duty to refresh and revitalize their bodies before returning to work. This helped in reducing their stress levels and to carry out their daily tasks more effectively. Some participants supported their claims with these statements,

> *I take a lot of rest when I am off duty. This gives me the chance to be refreshed and ready for the next day’s job at the hospital (Female, 40’s).*
>
> *The work is tedious so the only time I get to rest is when I am off duty (Male, 40’s).*
>
> *Resting is good and it serves a great purpose for me, especially when I am stressed at work. I usually will rest when I am off duty and that makes me feel better (Female, 30’s).*

#### Provision of accurate information to patients

To reduce the problem of con-compliance or non-collaborative patients, the FHWs interviewed stated that they tried to provide detailed and accurate information to the patients who tested positive to the virus. This was especially necessary for people who did not show any COVID-19 symptoms at the time. According to the participants, engaging in open and detailed communication helped in building and enhancing trust between them and their patients, calmed down the patients’ fears and anxieties, and ultimately, encouraged the patients to adhere to treatment and to follow safety protocols. Some participants captured these in the following statements.

> *Patients have the right to information, so providing them information to answer their questions must be right. One of the things I do is to ensure that I have accurate information about whatever the patient is asking (Female, 40’s).*
>
> *Patients ask a lot of question especially when they are isolated and does not display any signs of the COVID-19. That is when I take the opportunity to educate them (Male, 40’s).*
>
> *Giving out the right information is key in this era because patients are already worried about the COVID-19 so giving them wrong information will rather cost you. So, it is better to be sure of the information (Male, 40’s).*

#### Maintaining self-control and calmness

The participants interviewed agreed that a certain level of calmness and self-control were necessary in mitigating most of the issues they faced as FHWs. They explained that maintaining some level of calmness and self-control as FHWs helped them to provide effective treatment and care to patients diagnosed with COVID-19. They reported that when they themselves were calm, their patients were assured and encouraged to remain calm when seeking answers to their questions or getting treatment. This was especially true with people dealing with the loss of a loved one or news of a COVID-19 death. Following are some viewpoints expressed.

> *I have had cases where patients approach me in a hash manner demanding answers to their questions. I have to remain calm to have control of the situation (Female, 30’s).*
>
> *Encouraging patients that it is well and all this too shall pass gives them hope and puts smiles on their faces (Male, 30’s).*
>
> *Remaining calm was my style of professional service. Sometimes, patients meet you in a bad manner but as a professional physician I need to keep my cool and address the situation at hand (Female, 40’s).*

#### Teamwork and self-motivation

The study participants emphasized the importance of working in a team and self-motivation as important strategies they used to mitigate some of the challenges they faced as FHWs. They highlighted self-motivation as key to fulfilling their daily tasks, amidst the many challenges they faced such as high stress levels and lack of resources. They again emphasized that failure to motivate one’s self could easily lead to stress and job burnout. Below are statements made in support of these.

> *One thing that kept me going was my team, we compensate and motivate each other when we are overwhelmed with cases of COVID-19 (Female, 40’s).*
>
> *I most of the time encourage myself that this too shall pass and that God will protect me for me to serve others (Male, 30’s).*
>
> *My colleagues support me each time I get exhausted. I also do the same for them (Female, 40’s).*

### RQ 4. Motivation to perform daily tasks

COVID-19 brought untold psychological, emotional and physical toll on individuals, communities and the world in general. It was particularly stressful and emotionally challenging period for health care workers dealing with the pandemic. In view of these, participants of this study were asked to share their motivations for continuing as frontline health workers during this pandemic. Below are their responses.

#### Incentives and remuneration packages

All the participants identified incentives instituted by the Ghanaian government to FHWs as a major motivating factor. These included a boost in salary, insurance packages, and tax waivers for all FHWs.

##### Salary boost

The participants acknowledged that the President of the Republic of Ghana rewarded all frontline health workers with additional 50% of their basic salary to their current salaries for three months. According to them, this incentivised and boosted their enthusiasm to work. Three of the participants said,

> *I was so excited about the allowance provision made by the government. I was encouraged by it to work harder (Female, 30’s).*
>
> *The 50% basic allowance was exciting to me, I felt happy and willing to work (Male Physician, 40’s).*
>
> *The allowance encouraged me to work (Female, 40’s).*

##### Insurance package

In addition to the salary boost, the participants identified insurance packages established by the government to safeguard the lives and health of all frontline workers. Frontline health workers were insured for a total sum of GHc350, 000 (approx. US$29,411) per worker and covers hospitalization or death from COVID. The insurance provisions were specifically set up for health workers who directly interacted with COVID-19 patients, as captured by one of the interviewees,

> *I have to acknowledge that part of the support I receive is insurance coverage as a frontline worker (Female, 30’s)*

However, the participants expressed their discontent with the government’s inability to honor its obligation to pay the premiums of the insurance coverage. Two of the participants who contracted the virus complained that they paid their own hospital bills despite the government’s promise to take care of them. They had this to say,

> *Insurance package was one of the support services given to frontline workers but this insurance package has not yet been paid and I am disappointed (Male, 40’s).*
>
> *Insurance coverage for frontline workers was part of the support services but it has not been paid (Female, 30’s).*

##### Tax waiver

The study participants revealed that FHWs received tax waivers and other forms of incentives as part of their services rewards. They received exemptions for taxes on certain imports and additional benefits in the form of food items, accommodation, and on-site counselling by clinical psychologists. These motivated them to work. These statements were offered in support of this claim.

> *All frontline workers had a tax-free order when importing goods (Male, 50’s).*
>
> *Food, accommodation, and clinical psychologist were on site for use by frontline workers, this is part of the support services (Female, 40’s).*
>
> *I was accommodated and given food regularly as part of my support service as a frontline worker (Female, 30’s).*

#### Satisfaction from provision of service

Another motivating factor identified by the FHWs interviewed came from the satisfaction they derived from providing quality care and treatment to their patients. They categorized quality of service they provided to patients into high-quality and low-quality. High-quality services were characterised by effective monitoring, adequate resources, strict adherence to safety protocols, availability of vaccines, and patient satisfaction, while the low-quality service was characterised by poor monitoring, lack of resources, non-adherence to safety protocols, and shortage of vaccines. Majority of the FHWs interviewed claimed that, despite the challenges they experienced, they strived to provide high-quality service to their patients. They again claimed that they were motivated to work when their patients responded well to treatment. Below are some of the excerpts.

> *As a doctor, I am delighted when I am able to provide quality services to patients with covid-19 including vaccine shots, quick lab reports, and providing patients with PPEs. We are motivate to do more when patients respond positively to treatment (Male, 50’s).*
>
> *I gave COVID-19 patients proper treatment including basic counselling, going for lab results, proper monitoring, and ensuring adherence to safety protocols. This gives me a sense of satisfaction and motivation, especially in this tough times (Female, 40’s).*

However, a few of the participants indicated that they sometimes got demoralized when they were unable to provide satisfactory care or treatment to their patients. They classified their care as low-quality and demoralizing, as captured by two of the interviewee’s statements.

> *Honestly the services that I provided were not good enough from my perspective. This is due to lack of resources like PPEs and Vaccines (Female, 40’s).*
>
> *I considered my service to COVID-19 patients as low quality because we lacked PPEs and other vital resources like Ventilators. This demoralises me sometimes (Female, 30’s).*

## Discussion

The COVID-19 pandemic affected frontline healthcare workers all over the world, including Ghana. The novel pandemic pushed health facilities and their workers to develop different strategies as they strive to perform their tasks, save lives and defeat the virus. This study revealed that FHWs in Ghana perceived their work as highly risky, with a palpable anxiety and fear of contracting COVID-19 while caring for infected patients. Their fear and anxiety stemmed from the roles they performed as frontline defenders against the virus and its spread. These findings are consistent with other studies (Savini et al., 2017; Salazar de Pablo et al., 2020; Rogers et al., 2020; Lai et al., 2020; Ashinyo et al., 2020; Heinzerling et al., 2020). For instance, Ashinyo et al. (2020) revealed that frontline workers faced an increased risk of infection. Similar studies in Guinea by Savini et al. (2019) underscored the high exposure of health workers to COVID-19 infection, particularly those working in treatment centers. Lai et al. (2020) also found significant distress, depression, anxiety and insomnia among Chinese healthcare workers during the early stages of the pandemic. Other scholars such as Salazar de Pablo et al. (2020) and Rogers et al. (2020) have reported incidence of anxiety and depressive symptoms among healthcare workers.

Similar to findings of Beyond Blue (2019), Letvak et al. (2022), Kerlin, McPeake, & Mikkelsen (2020), Arpacioglu et al. (2020), Chen et al. (2020), Lai et al. (2020), and Wang et al. (2020), this study also revealed a range of challenges experienced by the FHWs interviewed, including burnout due to work overload, high incidence of stress and anxiety, difficulty balancing work and family lives, detachment from loved ones, sleep deprivation, and challenges related to the inadequate or lack of resources such as personal protective equipment.

However, contrary to findings of other studies, this research revealed a disturbing phenomenon whereby some Ghanaians, mostly wealthy people, who tested positive of the Covid-19 virus attempted to offer monetary enticements to physicians and lab technicians to change their positive test results to negative results to avoid quarantine or isolation. Others offered money to supervisors of isolation centers to help them leave the centers prematurely even though they still tested positive. This behaviour can be attributed to the normalization of bribery in the Ghanaian society, where it is expected for someone to pay bribe in order to get what they want done.

Frontline healthcare workers all over the world developed different strategies to help them perform their tasks during the COVID-19 pandemic. The experiences of the Ghanaian FHWs interviewed were no different. Similar to findings by Elkholy et al. (2020) and Feingold et al. (2021), this study revealed that FHWs took a lot of rest, sought psychological support from superiors or professionals, engaged in self-motivation, and constantly assured themselves of making positive impact in the lives of their patients. They also resorted to the use of technology, especially social media tools to bridge the communication and physical separation gap from their family members. In fact, this study highlights the importance of professional and psychological support received by the FHWs in coping with challenges they faced as first line defenders of the COVID-19 pandemic. These findings align with the recommendations by Maunder et al. (2013), Ofori et al. (2021) and Walton et al. (2020) that healthcare workers should engage the services of on-site psychologists or psychiatrists at their work places. Walton et al. (2020) went further to suggest the use of remote psychological support through phone, Zoom, or Skype as an effective alternative for providing support to frontline healthcare workers.

Unlike other studies, another unique observation of this study was the importance of the incentive packages created by the Ghana government to the frontline health workers. It seems that the majority of the FHWs interviewed were attracted to the incentives, which also motivated them to stay on the job regardless of the risk, stress, and other unsurmountable challenges they encountered. For example, the interviewees commended the government for increasing their salaries by an additional 50 percent and instituting insurance policies (although the government did not pay the premiums for most of the workers) and tax waivers as part of their support systems. This financial support system follows the suggestion by Cherish et al. (2020) that a risk allowance could serve as a strategy to motivate and retain healthcare workers during health crises. They also cautioned that such allowances might lead to jealousy among other staff who continue to care for non-COVID-19 patients and without those benefits.

## Conclusion

This study sought to investigate the experiences of frontline health workers of COVID-19 in Ghana. It relied on in-depth interviews of 32 FHWs drawn from three regions of Ghana to understand their experiences, challenges, strategies and motivations as they perform their duties. The findings overall provide useful insights into the work life of the frontline health worker. Although most of the findings of this study are consistent with observations and conclusions made by other researchers worldwide, two findings are worth noting as they seem unique to the experiences of FHWs in Ghana. The first is the monetary incentives offered by rich patients, their families or friends to the FHWs to act contrary to prescribed protocols for fighting the COVID-19 pandemic. This practice has a wider implication for delivery of effective care during pandemic situations, especially relating to the need for salary and incentive adjustments for FHWs, effective monitoring and enforcement of safety protocols, and institution of legal actions for violations of the protocols. The other finding worth noting is the importance of motivating healthcare workers, especially during serious health crisis and pandemics. In this study it was evident that the majority of the health workers (both volunteers and non-volunteers) were drawn to serve on the frontline of the fight against the COVID-19 virus because of the financial incentives. Unfortunately, as complained by the study participants, some of the financial incentives have not been honoured by the government. This can have serious implications for attracting and retaining such workers during future pandemics.

In conclusion, this study contributes new knowledge to this subject matter and provides useful information to guide future practical and policy guidelines for engaging frontline health workers during health emergencies and pandemics. A few recommendations are worth considering. Future studies should be expanded in scope to quantify the impact of the pandemic on the experiences of FHWs. This will help present a more complete and generalized understanding of the experiences of frontline health workers in the delivery of healthcare during pandemics. Governments faced with pandemics or health disasters should give adequate attention to the holistic welfare of the frontline health workers as well as provide a comprehensive and clear policy and legal frameworks to guide their work. Provisions such as adequate protective resources, support centers for psychological counselling, and establishment of financial and other forms of incentives are critical in attracting, motivating and retaining efficient staff. These will help reduce their fears, anxieties, stress and burnouts associated with the job. At the individual level, it is recommended that FHWs take mandatory breaks during work, practice mindfulness and other stress management exercises, and seek the needed support and counselling when working in pandemic situations.

### Public Health Implications

From a public health perspective, more emphasis must be placed on health education to improve public knowledge, awareness and understanding of pandemics and why it is important to adhere strictly to safety protocols and comply with treatment. This can positively impact people’s attitude and actions during health emergencies and curb bad practices that potentially can help spread the diseases, such as the offer of monetary incentives to frontline health workers to act contrary to established safety protocols and professionalism. Health emergencies and pandemics are bound to occur one way or another. Incentives motivate people to work harder to produce positive outcomes. However, some of the financial incentives were not honored and has still not been honored by the government, as indicated by some of the participants. This can have serious implications for attracting and retaining frontline health workers during health emergencies or pandemics. Our survival does not depend on whether they occur or not. It is what we do as health professionals, governments and the public during these times that makes the difference.

## Data Availability

All relevant data are within the manuscript and its Supporting Information files.

## Acknowledgments

We are deeply grateful to all the frontline health workers across Ghana who participated in this study. Your courage, resilience, and unwavering dedication in the face of the COVID-19 pandemic not only inspired this research but also shaped its findings in the most profound way. Thank you for sharing your stories with such honesty and openness.

Our sincere appreciation goes to the Ghana Health Service for granting ethical clearance and supporting access to healthcare facilities for data collection. We also acknowledge the assistance of the health directors, administrators, and staff at the various hospitals and clinics across the Greater Accra, Ashanti, and Western Regions, who facilitated this study’s successful implementation.

We are equally thankful to our families and friends for their continuous encouragement and support during the course of this research. Above all, we thank God for the strength, insight, and protection granted to us during this work.

## Notes

### Competing Interest Statement

The authors have declared no competing interest.

### Funding Statement

The author(s) received no specific funding for this work.

### Author Declarations

Ethics Review Committee of the Ghana Health Service (GHS) GHS-ERC:013/04/22

